# Forecasting COVID-19 cases using Machine Learning models

**DOI:** 10.1101/2020.07.02.20145474

**Authors:** Yuan Tian, Ishika Luthra, Xi Zhang

**Affiliations:** 33rd Conference on Neural Information Processing Systems (NeurIPS 2019), Vancouver, Canada

## Abstract

As of April 26, 2020, more than 2,994,958 cases of COVID-19 infection have been confirmed globally, raising a challenging public health issue. A predictive model of the disease would help allocate medical resources and determine social distancing measures more efficiently. In this paper, we gathered case data from Jan 22, 2020 to April 14 for 6 countries to compare different models’ proficiency in COVID-19 cases prediction. We assessed the performance of 3 machine learning models including hidden Markov chain model (HMM), hierarchical Bayes model, and long-short-term-memory model (LSTM) using the root-mean-square error (RMSE). The LSTM model had the consistently smallest prediction error rates for tracking the dynamics of incidents cases in 4 countries. In contrast, hierarchical Bayes model provided the most realistic prediction with the capability of identifying a plateau point in the incidents growth curve.

## 1 Introduction

The COVID-19 pandemic has spread devastatingly throughout the world, causing immense loss of life and economic hardships. It has played a toll in all aspects of human life, and become a great burden on hospitals and healthcare professionals. A key question that arises is how would the spread of the virus evolve, and how many people will get infected with this virus. The trend of confirmed cases varies greatly on the country studied. As well, there is great heterogeneity in response curves and determining when the virus has plateaued can affect social distancing measures. Forecasting the infection can also help with analysis of hospital resources that will be required. It can help countries prepare for the future and allocate their time and money. The goal of this paper is to predict the confirmed infection counts 5 days into the future. We aim to address the challenge in the novel, sparse and noisy data.

The known biological understanding of SARS/MERS was used as a basis for studying COVID-19, but this may not necessarily be reliable (Chinazzi et al. [2020], Peeri et al. [2020]). Recent studies (Read et al. [2020], Tang et al. [2020], Wu et al. [2020]) categorize people into the following four groups: susceptible (S), exposed (E), infected (I), and resistant (R) using a SEIR model. These methods hold strong assumptions about how the virus is transmitted, progresses in a patient and what people are most susceptible. When studying a novel outbreak it is best to not hold such biases to not skew results. Dandekar & Barbastathis, 2020 Dandekar and Barbastathis [2020] trained a Neural Network to model the spread of infection allowing for non-linear relationships to be observed which can occur once a quarantine is in place.

We compared three machine learning models to predict the number of confirmed infections in different countries. We identified LSTM model as the optimal model with lowest RMSE values. We also determined that hierarchical Bayes was the most realistic one in modelling the trend of the infection. It was able to find a plateau point at which the infection would stop growing in the population, a trend that was observed in past virus outbreaks.

## 2 Related Work

### 2.1 RNN-LSTM prediction of hand-foot-mouth disease

Gu et al., (2019) used weekly normalized hand-foot-mouth disease (HFMD) cases with meteorological data from 14 regions in Guangxi province, China to build a LSTM neural network for predicting new cases that arise. There are 14 factors in meteorological data, and 90 weeks of HFMD data available. The model we propose is slightly different from theirs in which we don’t have access to regional temperatures specific to each country, and it may not be very meaningful for larger countries. Nevertheless, both of our datasets are small. They only have data of 90 time points from each region and we have 84 time points from each country. Additionally, both models will be region-specific instead of one-size-fits-all [Gu et al., 2019].

### 2.2 Degune Forcast Model for Provinces in China

Guo et al.,(2017) Guo et al. [2017] trained several machine learning models including support vector regression model, step-down linear regression model, gradient boosted regression tree model, negative binomial regression model, least absolute shrinkage and selection operator linear regression model and generalized additive model. They used weekly dengue cases, Baidu search queries and climate factors as input features to predict the infected cases for 6 provinces in China. Performance from all models were assessed and validated through RMSE among different provinces and different forecasting windows. However, this study did not consider the dependency between different provinces’ data. We will address this limitation in our study by applying hierarchical Bayes algorithm.

### 2.3 The analysis of hospital infection data using hidden Markov models

Cooper, (2004) Cooper [2004] used a structured and un-structured hidden Markov model (HMM) to predict the spread of infection within hospitals. Understanding how infections are transmitted between patient-to-patient and even patient-to-staff can help protect those coming into the hospital and overall decrease the healthcare burden. The paper stated that the sparsity of time series data would be able to be handled using a HMM. A Poisson Distribution was used as the underlying distribution and 3 pathogens studied over approx. Forty months were used to evaluate their model. This paper’s model was very different from the one we proposed because it is modelling a very specific environment. The spread of infection in a hospital would be much quicker than in a regular community and patients coming in may already be immunocompromised. A limitation in both their study and ours is that using a HMM you do not take an individual patient’s susceptibility into account.

## 3 Motivation and Justification

### 3.1 Prediction of COVID-19 progression

We gathered the COVID-19 cases dataset from the Official Johns Hopkins University COVID github repository [Dong et al., 2020]. The record started from January 22nd and the cutoff date used by the models is April 14th, which consisted of 84 days in total. The cases recorded were confirmed clinically as well as cases confirmed by laboratory tests. It should be noted that clinically confirmed cases are more presumptive. To simplify the training, only confirmed cases were used. There were a total of 173 countries’ data available. Nevertheless, most countries have more than 30 days of zero cases, making the data sparse. A small subset of countries, however, were affected early on and have less than 30 non-zero data points. As well, the number of confirmed cases was expected to be limited by the speed and capability of testing for each country. In the end, the sparsity and noise problem would need to be addressed by the model.

### 3.2 Methods Summary

#### 3.2.1 Hierarchical Bayes Model

The proposed prediction problem contains data that is not from independent and identically distribution (IID). Since data points are IID only if they are from the same country, a hierarchical Bayes model was employed herein due to its relaxing IID assumption. Hyperpriors will be defined in this model, and it will introduce dependency between different countries’ sub models. Therefore, data-rich countries can inform posterior for data-poor countries.

#### 3.2.2 Long-Short-Term-Memory

LSTM was proposed as a method to be tested due to its strength in modelling time-series data. In contrast to other statistical models such as hierarchical Bayes, LSTM is naive and has no prior assumptions about statistical distribution and is suitable for novel datasets. The number of recurrent layers as well as number of hidden states features were selected from optimizing with validation sets specific to each model. The forget gates in LSTM corrects the vanishing gradient problem that can happen in deep RNN and can be updated at each time point, making LSTM suitable for modelling long sequences. This is beneficial for modelling COVID-19 as the magnitude of the change in data is relative to the stage of viral spread. As there is not a lot of data available for each country, Gaussian noise is added to the training process to augment the data and also to make the model more robust to noise in data and thus more generalizable.

#### 3.2.3 Hidden Markov Model

We decided to fit a HMM model to our COVID-19 data set as they have previously shown utility in modelling biological signal and correlations between events [Yoon, 2009]. They are able to capture the effect of an underlying state (may not be observable) in the signal that is being observed. In our case the signal being observed was the number of confirmed COVID-19 infections. Such modelling is usefully for studying spread of infections as the hidden states may directly correspond to when the virus is spreading (could be broken down further) and when it has plateaued. The initial state probability, transition probability, emission probability and number of hidden states are the model’s hyper parameters.

## 4 Experiments

### 4.1 Data processing

Daily COVID-19 infected cases for 173 countries from 1/22/2020 to 4/14/2020 were extracted from 2019 Novel Coronavirus COVID-19 (2019-nCoV) Data Repository by Johns Hopkins CSSE (data available: https://github.com/CSSEGISandData/COVID-19). The data was normalized per million of the country population data extracted from United Nations to prevent underflow. We compared RMSE of the last 5 days prediction (4/9-4/14) for certain chosen countries that have less than a month of zero cases among different 3 countries. Due to computation capacity limitation, only data in 6 countries including South Korea, Italy, United State, Taiwan, Japan, and Germany was used in training and validation.

### 4.2 Hidden Markov Model

Python version 3.7.4 and the hmmlearn library, version 0.2.3 were used to train and sample from the model. The three parameters we tuned in this model were: n_components (hidden states), covariance_type (covariance matrix) and n_iter (iterations).

#### 4.2.1 Method

n_iter was manually chosen to be 1000. The covariance_type was set to be a full covariance matrix as it led to the best results. To chose the number of hidden states we used the log probability of the model at different values and the RMSE for one country to predict the full 84 days, we also manually compared the graphs to confirm the correct trend was being capture. The results can be seen in the appendix section 6.1 Table 4. We chose 20 hidden components for our model because it had a relatively high log probability compared to the previous values tested and a low RMSE. This also meant that our model wasn’t too complicated.

Due to the noisy nature of the samples we also tried sampling over various runs and taking the mean over the samples at a specific time point. This led to a cleaner curve and decreased the RMSE for almost all countries so we used this approach. Graphs showing not averaged single sample case and table comparing RMSE can be found in the Appendix Section 6.2.

#### 4.2.2 Results

Once the model’s hyperparameters had been chosen, we trained the model on the first 79 days and used the model to sample the full 84 day dataset for 6 countries. The results can be seen below in Fig 1 and Table 1. Certain countries performed better than others but the general curve seemed to be captured through them all.

**Table 1:** HMM RMSE for Germany, Italy, United States, Taiwan, Japan, and South Korea for the averaged sampling over 100 samples predicted for 5 days.

|  | Germany | Italy | US | Taiwan | Japan | Korea, South | Average |
| --- | --- | --- | --- | --- | --- | --- | --- |
| RMSE | 284.72 | 436.86 | 324.61 | 1.99 | 7.42 | 78.15 | 188.96 |

**Figure 1:**
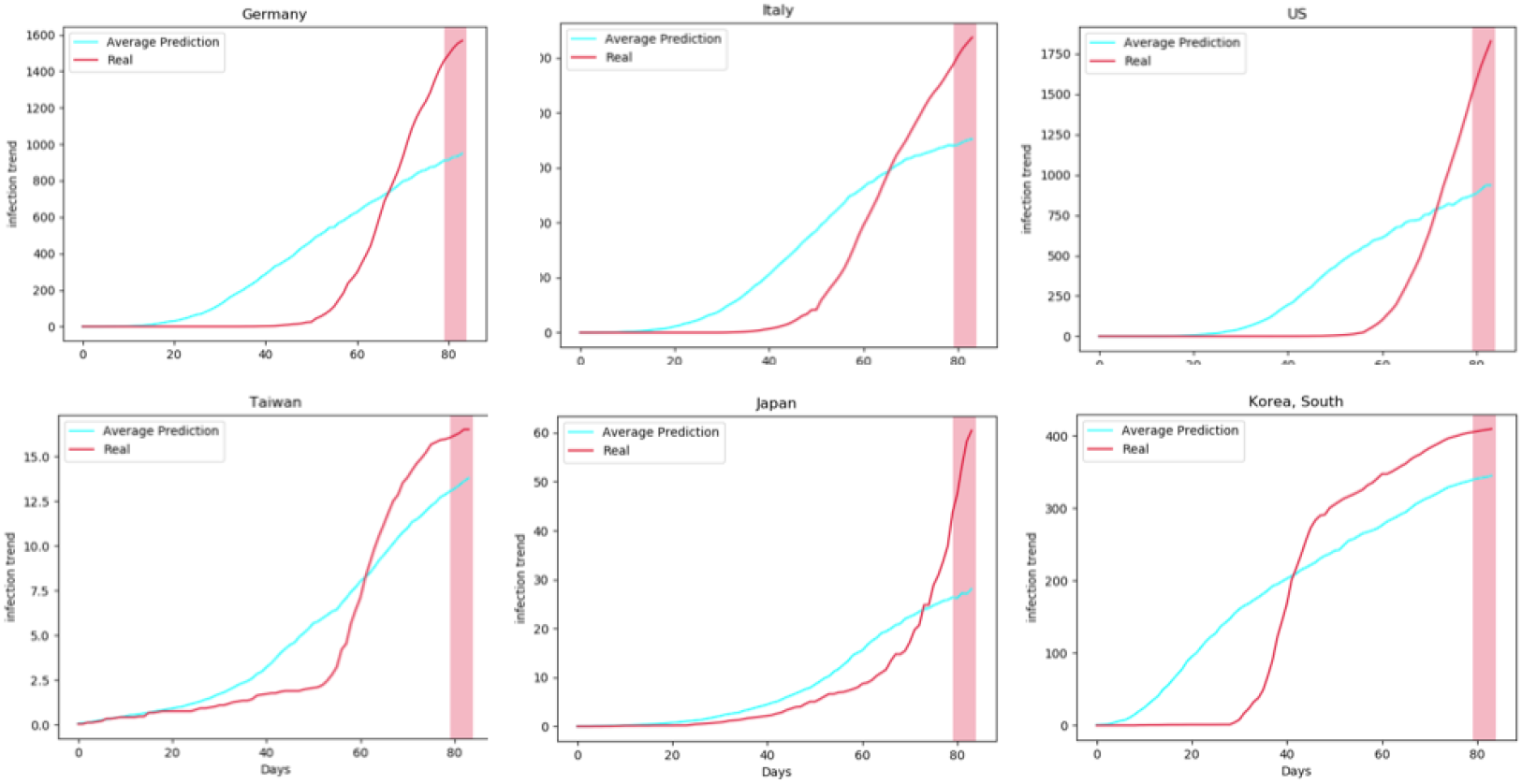
HMM Model Averaged over 100 samples for the 6 countries compared. The x-axis is number of days and the y-axis is number of confirmed cases normalized for each country.

### 4.3 Hierarchical Bayes Model

The structure of the hierarchical Bayes model consisted of parameters, priors and hyper priors. We assumed data points were IID given the country. Python 3.7 and PyMC3 library were used for implementing the model.

#### 4.3.1 Method

In this model, we employed the generalized logistic function to model the infected cases growth curve for each country j.

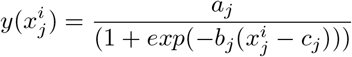

where 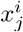 is the *i*th day after the first day that there were confirmed cases reported in this country j, and *a*_*j*_, *b*_*j*_, *c*_*j*_ are the model parameters for country j.

We assumed that *a*_*j*_, *b*_*j*_, *c*_*j*_ for each country j were drawn from common prior,

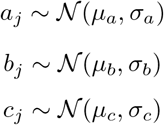

and we also introduced hyperpriors for each prior such that we can model the dependency between each country’s sub model.

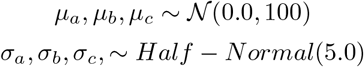

#### 4.3.2 Results

Table 2 shows the RMSE for the prediction of testing period in each country. The average RMSE is 92.10 in units of cases per millionth of people.

**Table 2:** Hierarchical Bayes Model RMSE for Germany, Italy, United States, Taiwan, Japan, and South Korea with respect to forecast window of 5 days.

|  | Germany | Italy | US | Taiwan | Japan | Korea, South | Average |
| --- | --- | --- | --- | --- | --- | --- | --- |
| RMSE | 73.9 | 226.22 | 184.29 | 18.28 | 12.87 | 37.04 | 92.10 |

Figure 2 shows the model’s performance for each country. The x-axis is the days after the first day that there were confirmed cases reported in this country, corresponding to the dates 22 Jan 2020 to 14 Apr 2020. In addition, for each country, data points with 0 confirmed case were removed from the training set. The mean model curve that uses the mean value of parameters among all sub models was also plotted in the figured as a baseline. The average RMSE in units of cases per millionth of people is around 92.10. As we can see hierarchical Bayes model tends to underestimate the confirmed cases.

**Figure 2:**
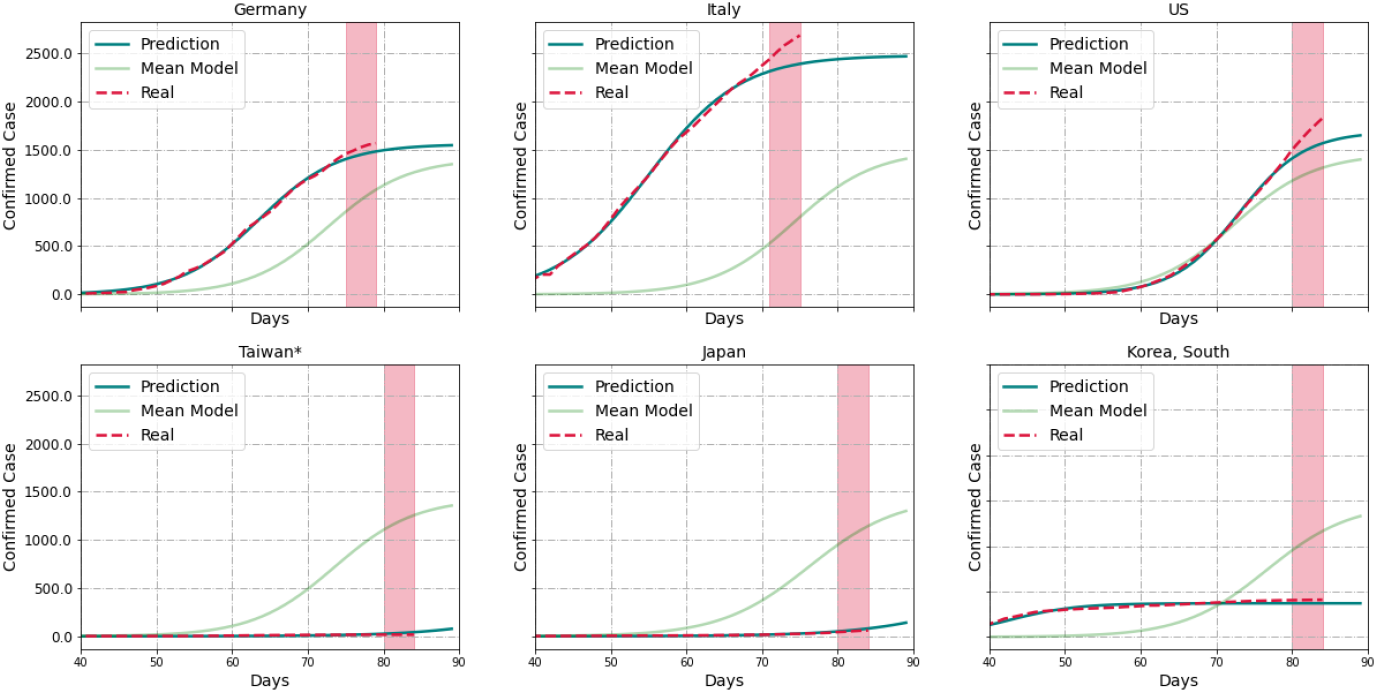
Hierarchical Bayes Model Performance for Germany, Italy, United States, Taiwan, Japan, and South Korea with Respect to Forecast Window of 5 Days. The x axis is the days after there were first confirmed cases reported in this country, and the y axis is the confirmed cases per million of the country population. The shaded region is the 5-day testing period.

### 4.4 Long-Short-Term-Memory RNN

The LSTM model was implemented via Pytorch v1.4.0, with model-specific optimized hyperparameters of number of hidden states, number of layers, learning rate and momentum of stochastic gradient descent (SGD) momentum.

The layers from LSTM applied the following transformation at input time t where *b* is bias, and *h* is hidden states:

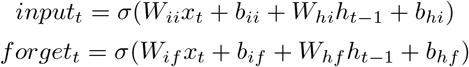

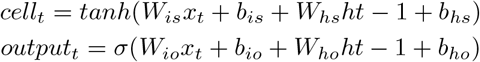

The hidden state is determined by the cell state, which consists of what is not forgotten from the previous cell state and the current input. As well, the hidden state is *tanh* transformed.

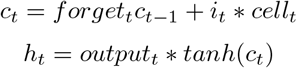

In order to have sizable training sets, twenty-five days prior to the 5 day testing sequence was used as inputs. The last 5 days was used for testing, and the last 5th to 10th day was used for validation. The rest of the data were used for training. To prevent learning mostly zero sequences, the data was only used after the first non-zero case date. As well, the data used to train the model was the change in number of confirmed cases instead of the number of cases to prevent the model from learning auto-correlation in the data. As the dataset size was still modest, Gaussian noise with variance of a thousandth of the max value in training set was added for data augmentation. Following the LSTM layers, a linear layer and ReLU was applied. ReLU was used to correct the LSTM model’s tendency in predicting negative values even when none of the training data contained negative values.

#### 4.4.1 LSTM prediction result

The result of the LSTM model is shown in table 3 and figure 3. As seen from the plot (higher resolution figure in appendix), the LSTM model tends to predict higher case numbers for all countries except for Japan. The average RMSE in units of cases per millionth of people is around 63.88.

**Table 3:** Long-Short-Term-Memory RMSE for the six chosen countries of the 5 day prediction. Unit is in case per millionth people.

|  | Germany | Italy | US | Taiwan | Japan | Korea, South | Average |
| --- | --- | --- | --- | --- | --- | --- | --- |
| LSTM RMSE | 74.98 | 258.65 | 27.61 | 1.10 | 9.91 | 11.04 | 63.88 |

**Figure 3:**
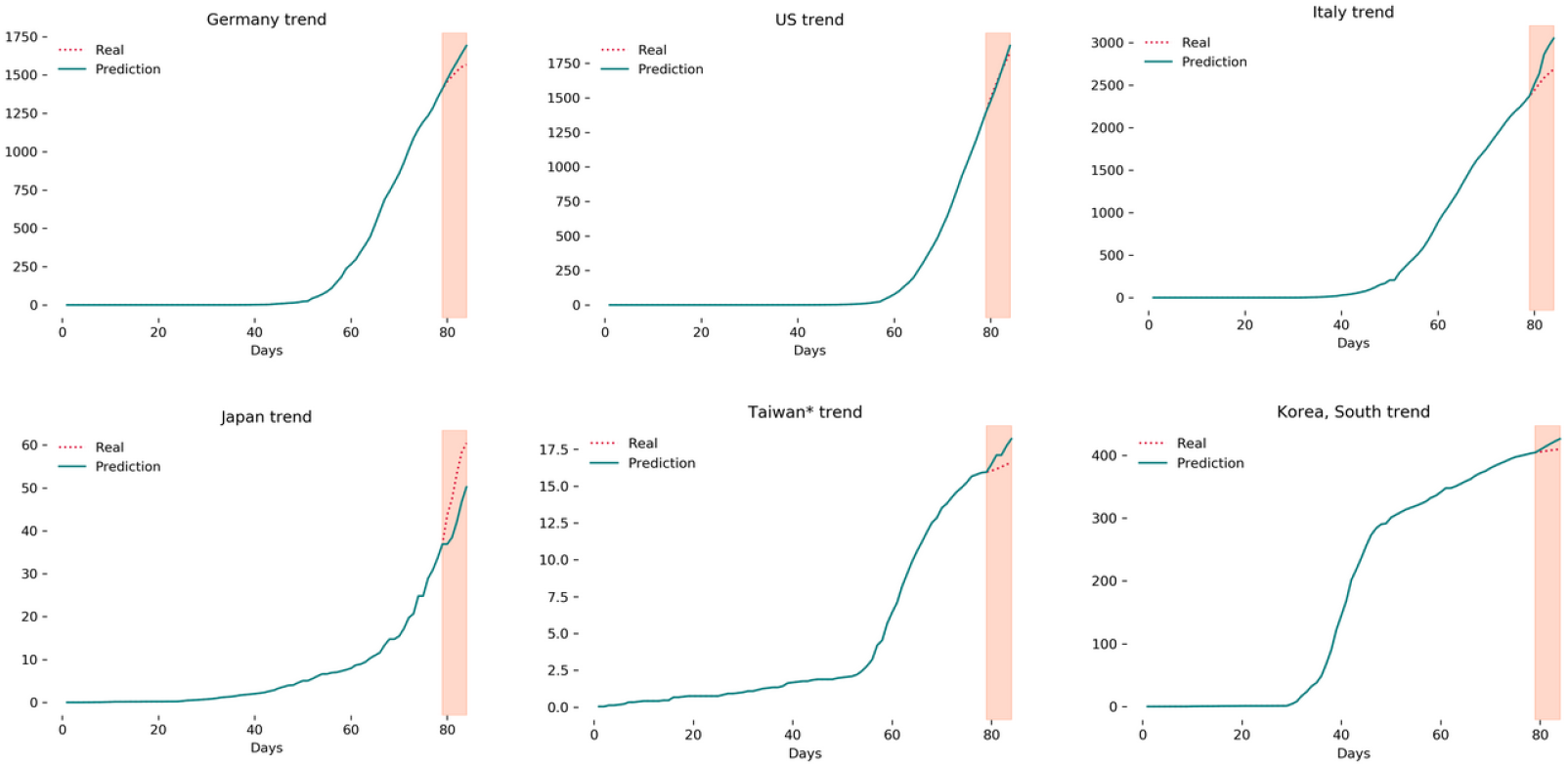
Long-Short-Term-Memory RNN prediction of COVID confirmed cases for the specified countries for 5 days normalized to the millionth of the population. The graph shows the projection in the shaded region with the teal line showing prediction and the crimson dotted line showing the real data. The high-resolution figure along with its zoomed-in projection can be seen in appendix

### 4.5 Comparison of Prediction Performance between Models

Figure 4 shows the comparison of prediction performance among HMM model, hierarchical Bayes model, and LSTM model by RMSE. As we can see, LSTM model has the lowest RMSE for most of the countries other than Italy, while HMM models performs poorly among most countries with much higher RMSE than other models. Hierarchical Bayes model has close RMSE values as LSTM model, and it even performs better than LSTM model for predicting confirmed cases in Italy and Germany. For the general trend of model prediction, hierarchical Bayes model and tends to overestimate the cases while HMM model and LSTM model tend to underestimate the number of cases. +

**Figure 4:**
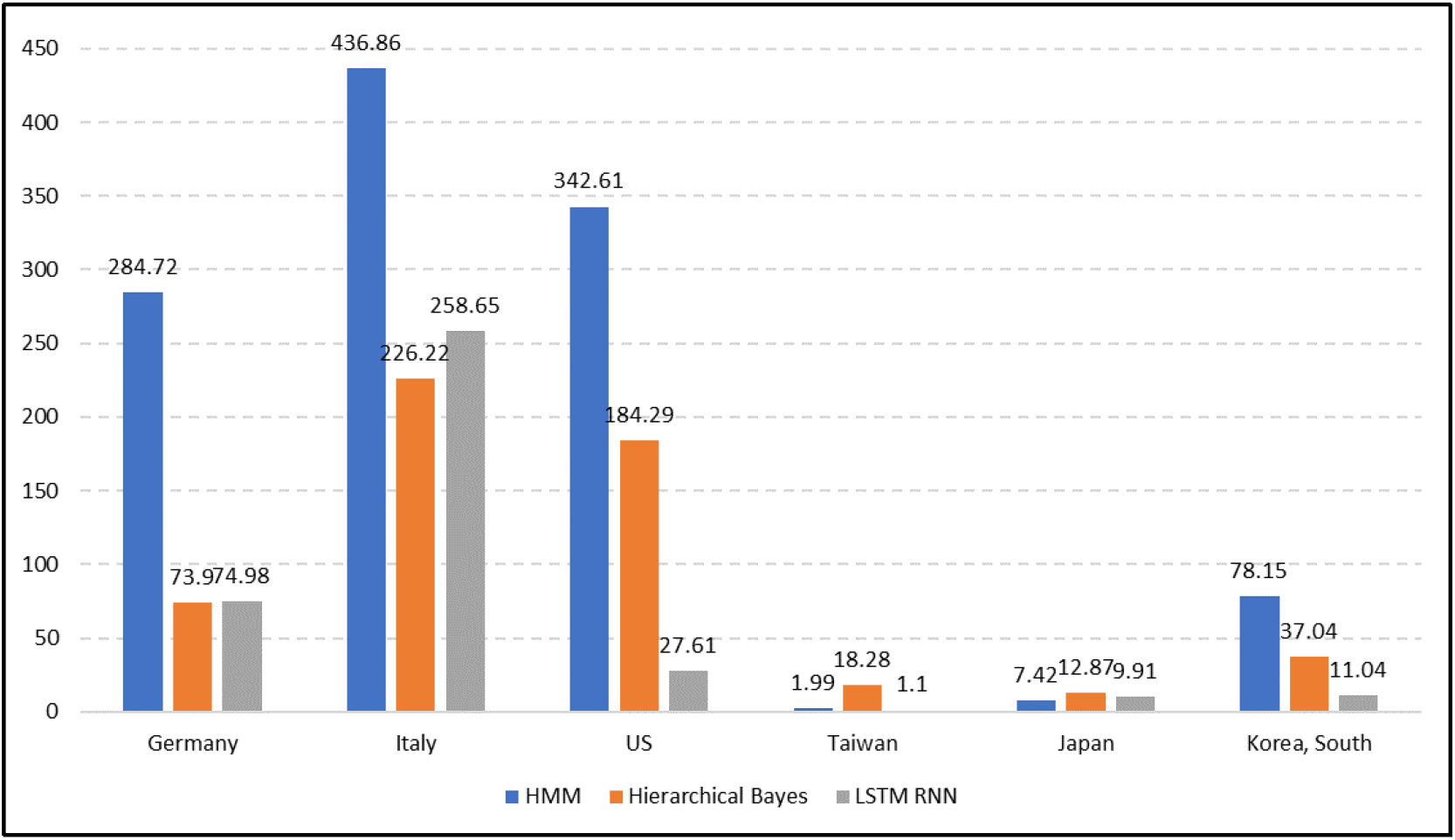
Comparison of Prediction Performance of the Models by RMSE. LSTM model performs best for most of countries including Germany, US, Taiwan, Japan, South Korea, while HMM performs the worst for most of countries.

## 5 Discussion and Future direction

This study demonstrated LSTM was an accurate model in predicting the COVID-19 epidemic trajectory for 6 selected countries. We thoroughly evaluated 3 machine learning models for COVID-19 confirmed cases prediction, and identify LSTM model as the optimal model that may help improve the surveillance for infectious disease. Even though LSTM model has the lowest RMSE for most of countries, hierarchical Bayes model performs better than LSTM for 2 countries. Therefore, both method should be considered while modeling the infectious disease outbreak.

As for HMM model, the forecasts were not able to correctly model the infection trend. The results were under extreme stochastic effects and the plots and RMSE varied greatly over different runs. Therefore, HMM seem to be very sensitive to initialization and may not be reliable in producing consistent results. With more time using a different underlying distribution (ex. Poisson) might lead more encouraging results.

For hierarchical Bayes model, on the one hand it considered dependency between data from different countries while other models could not, on the other hand, we assumed generalized logistic function to model the growth curve in this study, which lacked the dependency between adjacent days and may fail when the assumption is false. In the future work, we will try to combine HMM and hierarchical Bayes to capture the connection in time series.

The LSTM model learned the long-term dependencies between data and was supposed to be able to tolerate noise in data as noise was added during the training process. The model had similar RMSE to hierarchical Bayes but it may be constrained by the training data as most countries have not had reached the plateau phase in the viral spread. In the future, transfer learning using other infectious disease data or data from countries affected early on may be used to initialize the weights of the model so the plateau phase is considered.

Overall, a caveat of our study is that none of the countries we studied reached a plateau in their infection count. To complete a holistic search of models, given more time, we should compare across more models, train on a larger data set, and increase the prediction window. Future work would also include integrating other factors that play a role in the spread of infection such as geographic location, population density, GDP, number of hospitals and doctors etc. In addition, applying ensemble method to average the results from LSTM model and hierarchical Bayes model may also provide a more accurate prediction model.

## Data Availability

The code used in the manuscript is available at: https://github.com/nZhangx/CPSC540_Covid

## 6 Appendix

### 6.1 LSTM performance

**Figure 5:**
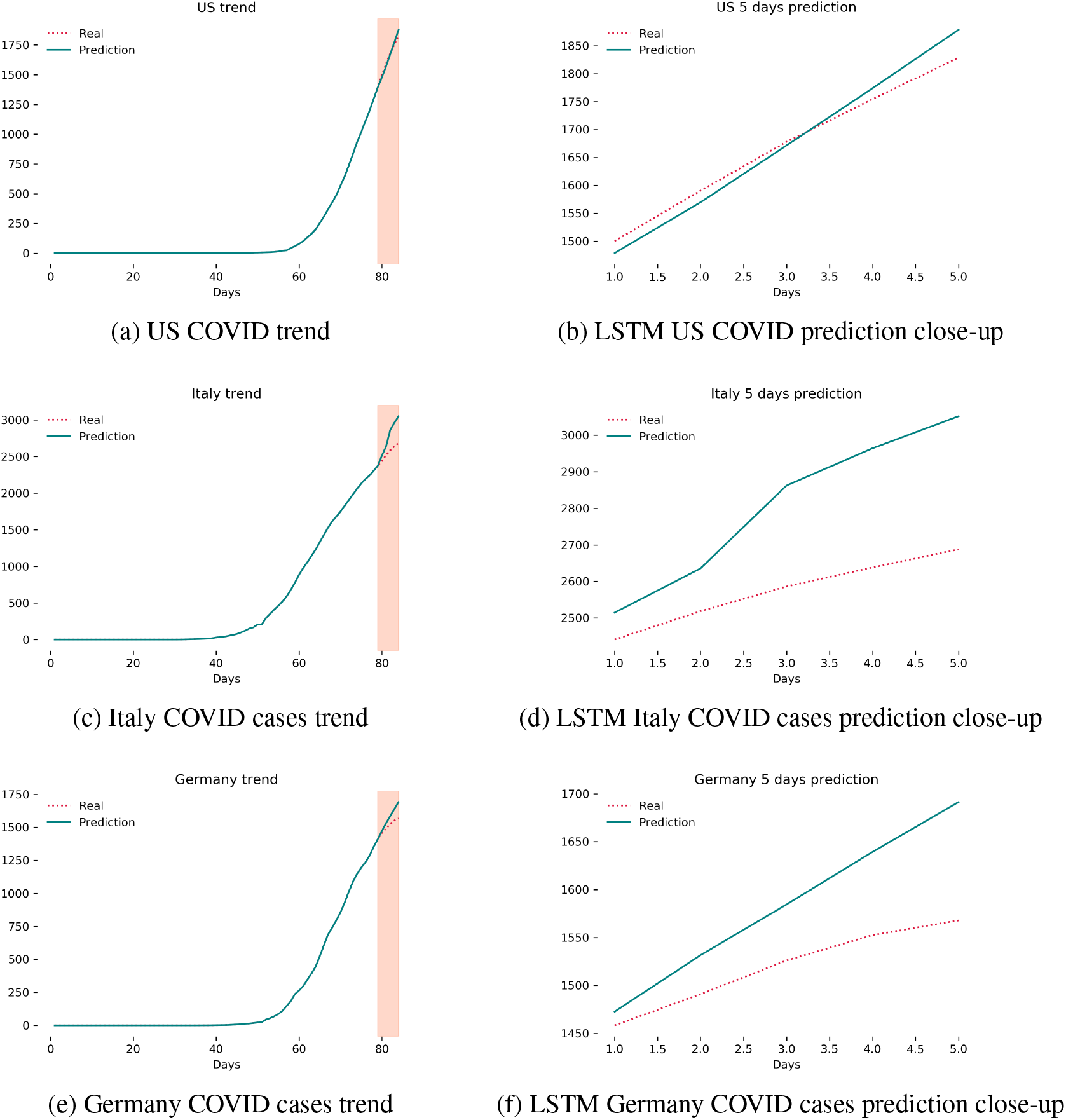
COVID-cases close-up

### 6.2 HMM performance

**Table 4:** RMSE and log probability of each model fitted for Italy with a varying number of hidden states. The log probability always increases as the model becomes more complicated. Future work would include taking into account how these values change of other countries and using a AIC/BIC approach to choose the number of hidden components.

| n_components | RMSE | log Probability |
| --- | --- | --- |
| 2 | 992.53 | -378.94 |
| 5 | 743.13 | -280.01 |
| 10 | 148.98 | -239.13 |
| 20 | 314.23 | -206.40 |
| 30 | 1069.78 | -105.06 |
| 40 | 839.24 | -7.45 |
| 50 | 420.53 | 37.60 |

**Table 5:** HMM RMSE for one sample for Germany, Italy, United States, Taiwan, Japan, and South Korea predicting 5 days.

|  | Germany | Italy | US | Taiwan | Japan | Korea, South | Average |
| --- | --- | --- | --- | --- | --- | --- | --- |
| RMSE | 667.22 | 452.76 | 118.28 | 4.58 | 9.47 | 279.71 | 255.34 |

**Figure 6:**
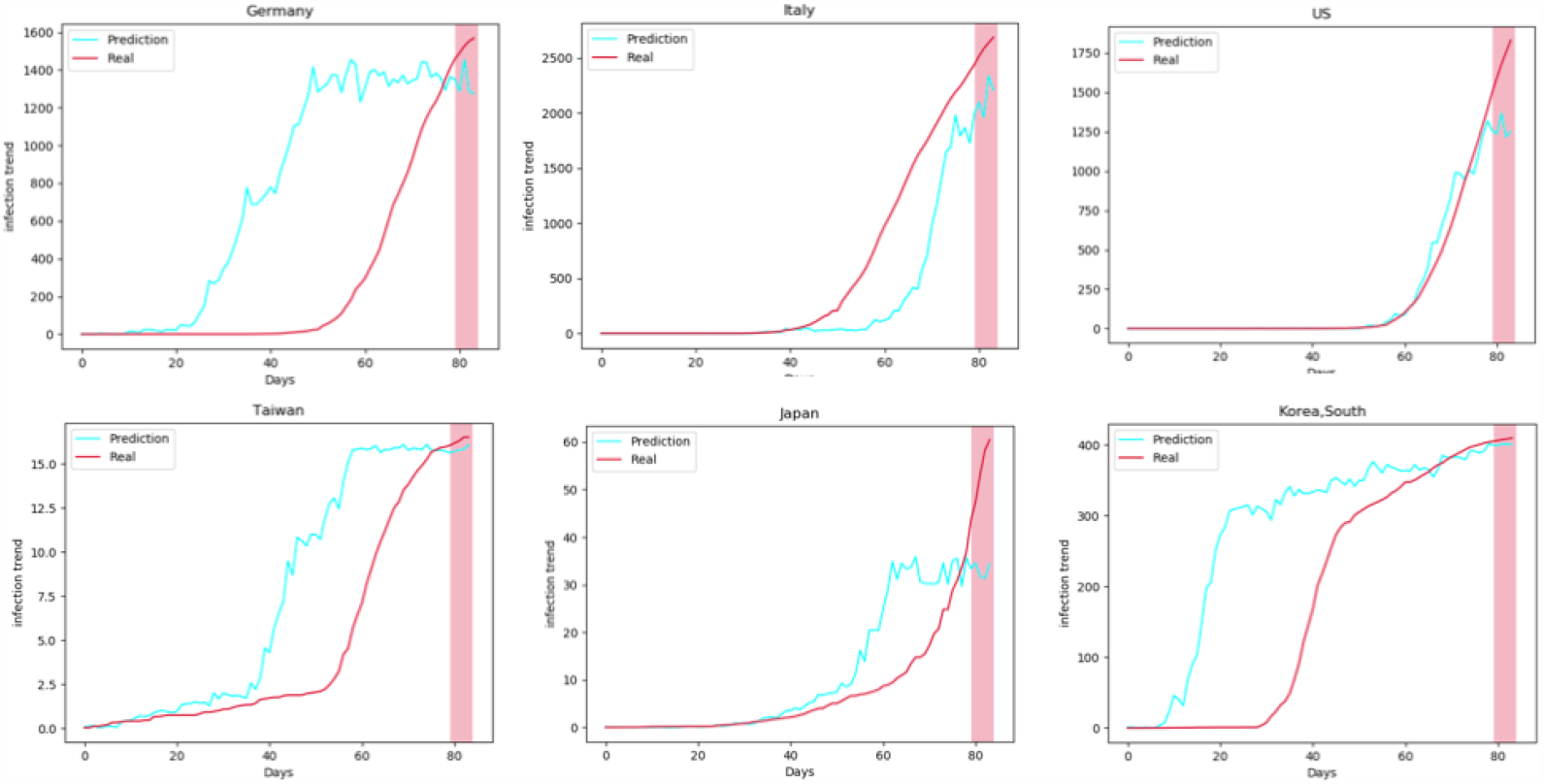
HMM Model for one samples for the 6 countries compared. The x-axis is number of days and the y-axis is number of confirmed cases normalized for each country.

